# Early human capital, the COVID-19 food-insecurity shock, and adult mental health: a 22-year cohort study in Ethiopia, India, and Peru

**DOI:** 10.64898/2026.09.02.26362122

**Authors:** Manuel Antonio Díaz Flores, William Miguel Jiménez Rivera, Gina Milagros Peña Huamán, Ursula Romero Molina

## Abstract

Mental disorders typically emerge before age 25, yet little is known about how childhood human capital and large economic shocks jointly shape mental health into adulthood in low-and middle-income countries (LMICs). We use seven rounds of the Young Lives study (2002–2024), which followed two birth cohorts in Ethiopia, India, Peru, and Vietnam, to examine (i) the long reach of early human capital, proxied by the height-for-age *z*-score in infancy; (ii) the medium-term association between household food insecurity during the COVID-19 pandemic and adult depressive (PHQ-8) and anxiety (GAD-7) symptoms measured at ages 22 and 29 in 2023–2024; and (iii) whether early human capital buffered the pandemic shock. We estimate value-added, latent-factor, dynamic panel, and mediation models with cluster-robust inference. Early height-for-age strongly predicts adult height (1.6 cm per standard deviation, *p <* 0.01) but not adult mental health or formal employment. Pandemic food insecurity predicts higher depressive (0.41 points, 0.12 standard deviations) and anxiety (0.41 points, 0.11 standard deviations) symptoms three to four years later (both *p <* 0.01); per step, the probability of clinically relevant anxiety (GAD-7 of 10 or more; base 7.6 percent) rises by 1.9 percentage points (*p* = 0.001) and of clinically relevant depression (PHQ-8 of 10 or more; base 5.6 percent) by 1.0 percentage point (*p* = 0.07). The gradient travels with the persistence of distress and of material hardship, is robust to attrition corrections and bounds, and its causal reading is explicitly bounded, since mental health was first measured during the pandemic itself. Exploratory analyses suggest the gradient differs across countries and may be larger in the older cohort, exposed at about age 26 rather than 19; these contrasts rest on few clusters and are not robust to multiple-comparison adjustment. Early human capital neither predicts adult mental health—a precise null of at most *±*0.03 standard deviations—nor moderates the gradient. Protecting household food security during aggregate shocks, especially for young adults, appears central to safeguarding mental health in LMICs.

## Introduction

Mental disorders are a leading cause of disability worldwide [1] and typically have their onset before age 25 [2], yet most longitudinal evidence on their early-life determinants comes from high-income settings. In low- and middle-income countries (LMICs), where the majority of the world’s young people live, the joint role of childhood human capital and large economic shocks in shaping mental health into adulthood remains poorly understood [3], largely because few cohorts have followed the same children from infancy into their twenties.

Two literatures bear on this question but have rarely been connected at the adult margin. The first, on the technology of skill formation [4–6], shows that human capital is produced cumulatively, with self-productivity and dynamic complementarity, and that early endowments—often proxied by height-for-age, a summary measure of early-life net nutrition [7]—have a long reach into later cognitive, educational, and labour-market outcomes [8–13]. The second documents the immediate mental-health toll of the COVID-19 pandemic in LMICs. Using the Young Lives COVID-19 phone survey, Porter et al. [14] and Favara et al. [15] showed that anxiety and depression rose during 2020 and were tightly linked to household food insecurity across four countries, and Hamadani et al. [16] documented the same food-insecurity channel in Bangladesh. What happened next—whether the pandemic-period deterioration was followed by a *medium-term* gradient once economies reopened, whether any such gradient depended on national context and on the life stage at which young people were hit, and whether children who entered the pandemic with more human capital proved more resilient—has not been studied, because the relevant adult data only became available with the seventh Young Lives round in 2023–2024.

This paper fills that gap. We combine the full seven-round Young Lives panel to test three linked propositions. First (P1), that early human capital has a long reach into adult outcomes measured at ages 22 and 29. Second (P2), that the pandemic food-insecurity shock is associated with worse adult mental health several years later. Third (P3)—the integrating hypothesis—that early human capital buffered the pandemic shock, as dynamic complementarity [4] would predict. Because the design observes two birth cohorts in several countries under a common protocol, we can also ask *where* and *for whom* the gradient is largest, comparing its size across countries, across cohorts exposed at different life stages, and between women and men—comparisons we label exploratory throughout, given the number of contrasts involved.

Our contribution is threefold. We extend the human-capital and pandemic mental-health literatures from childhood and the acute pandemic period, respectively, to young-adult outcomes newly observed in 2023–2024; we provide, to the best of our knowledge, an early test of the resilience hypothesis with accumulated human capital (rather than a single programme) in LMICs; and we document suggestive evidence that the mental-health legacy of COVID-19 varies with national context and with the life stage at which the shock arrived.

## Materials and methods

### Ethics statement

This study is a secondary analysis of fully anonymised, publicly archived data collected by the Young Lives study, which obtained ethical approval from the University of Oxford Central University Research Ethics Committee and from country-level committees. Informed consent—written or, where literacy was limited, documented oral consent—was obtained from caregivers and participants at every round. The anonymised data were accessed through the UK Data Service (study number 9543); no additional ethical approval was required for this analysis.

### The Young Lives cohort

Young Lives is a longitudinal study that has followed two birth cohorts—a Younger Cohort born around 2001 and an Older Cohort born around 1994—in Ethiopia, India (the states of Andhra Pradesh and Telangana), Peru, and Vietnam since 2002 [17]. We use the Round 1–7 constructed files (UK Data Service study 9543), which harmonise variables across the five in-person rounds (2002–2016), the 2020–2021 COVID-19 phone survey (Round 6), and the in-person Round 7 (2023–2024). At Round 7 the Younger and Older Cohorts were approximately 22 and 29 years old; during the 2020 phone survey they were approximately 19 and 26. Round 7 was not fielded in Vietnam; adult outcomes are therefore available for Ethiopia, India, and Peru, while Vietnam contributes to the childhood and pandemic-period measures. The sample flow from enrolment to the analytic sample is as follows: of the 11,784 children ever enrolled across the four countries, 11,586 have valid Round-1 anthropometry; 8,500 were reached by the Round-6 phone survey with the food-security module; 6,754 have an adult PHQ-8 score at Round 7 (possible only in the three countries where the round was fielded); and the COVID specification uses the 5,135 participants with complete data on the exposure, the outcome, and the full predetermined vector. Missing data are handled by listwise deletion within each model and no imputation is performed, so each table reports its own estimation sample. The study is reported in accordance with the STROBE guidelines for observational studies (S2 Checklist).

### Outcomes

Our primary outcome is depressive symptoms in adulthood, measured with the eight-item Patient Health Questionnaire (PHQ-8, range 0–24) [18]; we also examine anxiety with the seven-item Generalized Anxiety Disorder scale (GAD-7, range 0–21) [19]. Secondary outcomes include formal employment (holding a written contract), adult height, and completed schooling. Anthropometric *z*-scores were cleaned using the WHO biological-plausibility flags, with *|z| >* 6 set to missing [20].

### Exposures and covariates

Early human capital is proxied by the height-for-age *z*-score (HAZ) at Round 1 (age one for the Younger Cohort). Pandemic exposure is captured by household food insecurity reported during Round 6 on a four-point scale (1 = always eats enough to 4 = frequently does not eat enough), taking each participant’s worst report across the phone-survey calls; the household economic-shock modules were not populated in the constructed Round 6 files, so food insecurity—the very channel highlighted in the acute-period literature [15]—is the available pandemic stressor. The predetermined covariate vector ***X****_i_* includes early stunting, the three Young Lives wealth sub-indices (housing quality, access to services, consumer durables), maternal and paternal education, sex, urban residence, a cumulative count of rounds with household shocks across childhood (0–5), motivated by cumulative-risk models of development [21], and cohort and country fixed effects. Because early stunting is the indicator 1[HAZ *< −*2], a deterministic transformation of the continuous score, specifications that include both amount to a linear spline with a knot at *−*2: the coefficient on the continuous score is then a slope away from the knot, not a total association. We therefore also report the key equations with the continuous score alone, and we read the stunting coefficients as spline segments rather than as a separate construct. The value-added specification additionally conditions on lagged pandemic distress, contemporaneous adult food insecurity, disability, and marital status. Table 1 reports descriptive statistics for all analysis variables.

**Table 1.** Descriptive statistics of the analytic sample.

|  | Mean | SD | Min | Max | N |
| --- | --- | --- | --- | --- | --- |
| Height-for-age z (R1) | −1.37 | 1.38 | −6.0 | 5.9 | 11586 |
| Early stunting (R1) | 0.30 | 0.46 | 0.0 | 1.0 | 11586 |
| Housing quality (R1) | 0.38 | 0.27 | 0.0 | 1.0 | 8777 |
| Services access (R1) | 0.50 | 0.34 | 0.0 | 1.0 | 8768 |
| Consumer durables (R1) | 0.19 | 0.19 | 0.0 | 1.0 | 8766 |
| Mother’s education (grades) | 4.44 | 4.81 | 0.0 | 30.0 | 11428 |
| Father’s education (grades) | 5.47 | 5.07 | 0.0 | 29.0 | 10367 |
| Female | 0.48 | 0.50 | 0.0 | 1.0 | 11784 |
| Urban | 0.37 | 0.48 | 0.0 | 1.0 | 11784 |
| Cumulative household shocks (0–5) | 3.14 | 1.33 | 0.0 | 5.0 | 11784 |
| Food insecurity, COVID (R6) | 1.80 | 0.58 | 1.0 | 4.0 | 8500 |
| PHQ-8 during COVID (R6) | 1.91 | 2.36 | 0.0 | 19.0 | 9224 |
| Food insecurity, adult (R7) | 1.90 | 0.58 | 1.0 | 4.0 | 7123 |
| Permanent disability (R7) | 0.02 | 0.14 | 0.0 | 1.0 | 6926 |
| Married/cohabiting (R7) | 0.30 | 0.46 | 0.0 | 1.0 | 7118 |
| PHQ-8 adult (R7) | 2.30 | 3.50 | 0.0 | 24.0 | 6754 |
| GAD-7 adult (R7) | 2.77 | 3.79 | 0.0 | 21.0 | 6927 |
| Written contract (R7) | 0.16 | 0.37 | 0.0 | 1.0 | 4442 |
| Grades completed (R7) | 10.12 | 2.35 | 0.0 | 12.0 | 7092 |
| Adult height, cm (R7) | 161.65 | 9.12 | 126.5 | 196.1 | 6844 |

R1, R6, and R7 denote Young Lives Rounds 1 (2002), 6 (2020–2021 phone survey), and 7 (2023–2024). Wealth sub-indices range from 0 to 1. Sample sizes vary because Round 7 was not fielded in Vietnam and because of item non-response. Adult height is subjected to adult plausibility limits (120–210 cm), which removes one implausible record.

Fig 1 shows the characteristic early growth faltering and partial recovery of height-for-age across the four countries, a pattern documented in the Young Lives anthropometric literature [9, 22].

**Fig 1.**
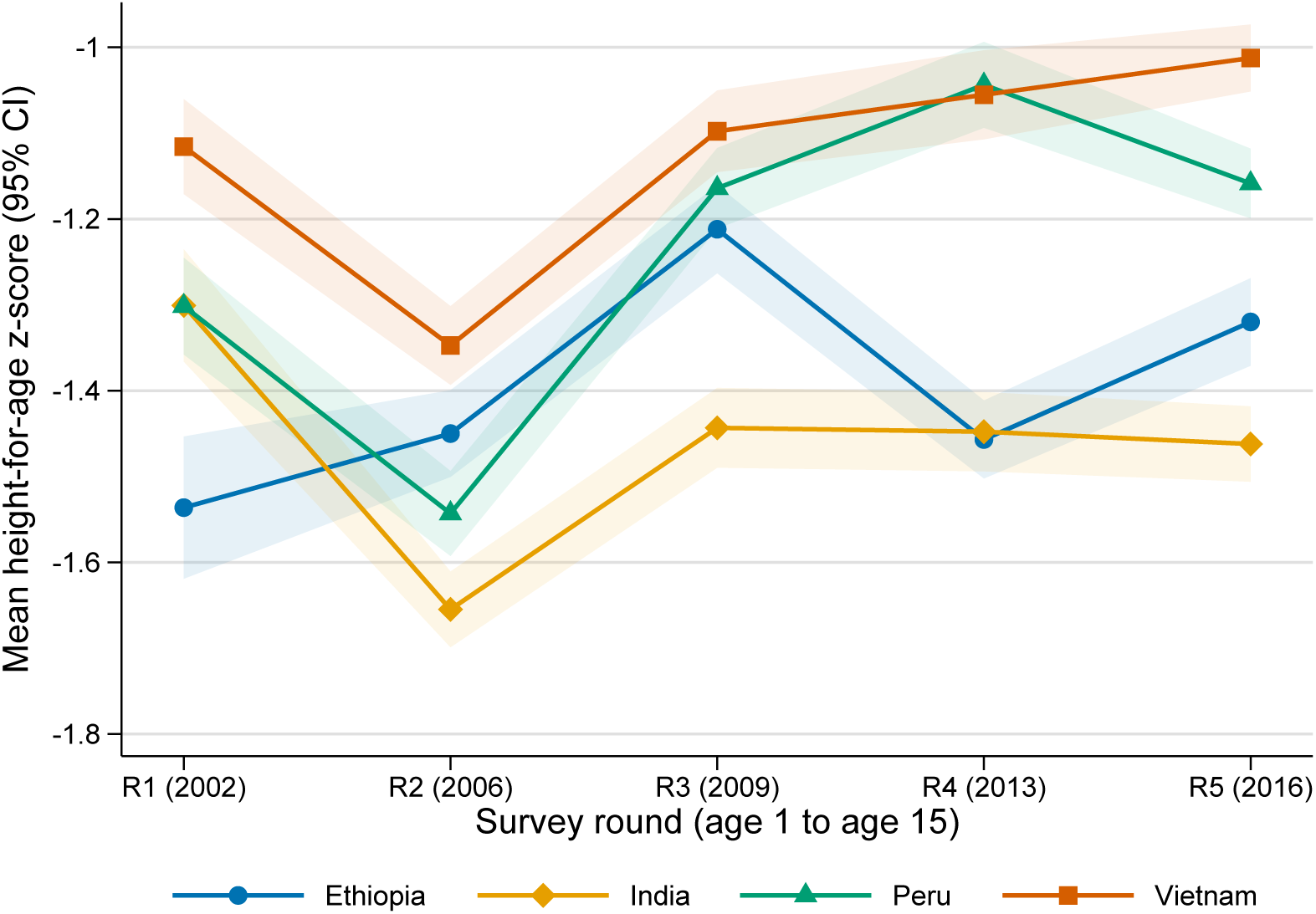
Growth faltering and partial recovery in the Younger Cohort. Mean height-for-age *z*-score by survey round and country for the Younger Cohort (Young Lives Rounds 1–5, ages 1 to 15), with 95% confidence bands around each country mean. Higher values indicate better cumulative net nutrition. *z*-scores are cleaned using WHO biological-plausibility flags.

### Statistical analysis

#### Production function and value-added models

We anchor the analysis in the cumulative human-capital production function [6, 23], in which an adult outcome *Y_i_* is the accumulated product of early endowments, inputs, and shocks:

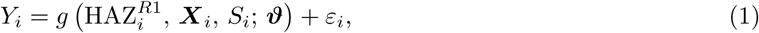

where 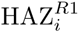 is early human capital, ***X****_i_* the predetermined covariate vector, and *S_i_* a vector of shocks. We approximate Eq (1) linearly. The baseline and COVID specifications regress adult depression on early human capital, the predetermined covariates, and—in the COVID specification—pandemic food insecurity:

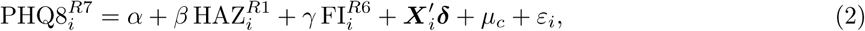

where 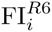 is Round-6 food insecurity, *µ_c_* are country fixed effects, and *γ* is the parameter of interest for proposition P2. Entering the ordinal 1–4 scale linearly imposes equidistant category steps; we therefore also estimate Eq (2) with the linear term replaced by category indicators, test the equidistance restriction with a Wald test, and additionally use the binary moderate/severe recoding (Appendix Table A7). Because the symptom scores are bounded counts with a large mass at zero, we further verify the linear results with Poisson quasi-maximum-likelihood models and with logit models for the clinical thresholds (PHQ-8 *≥* 10, GAD-7 *≥* 10), and we report effect sizes both in standard-deviation units and as changes in the probability of exceeding the clinical threshold (Appendix Table A9). The value-added specification augments Eq (2) with the lagged pandemic-period symptom score and contemporaneous adult correlates, following the conditional-on-lagged-outcome logic of Todd and Wolpin [23]:

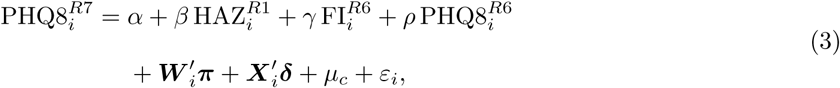

where ***W*** *_i_* collects adult food insecurity, disability, and marital status. The same specifications are estimated for anxiety (GAD-7). The division of labour between the two equations should be explicit. Eq (2), which conditions only on predetermined covariates, is the *primary* specification: the magnitude of the pandemic gradient *γ* should be read from it, and only from it. The added regressors in Eq (3)—lagged pandemic distress and contemporaneous adult circumstances—are measured *after* the exposures of interest and are plausibly on their causal pathway. Conditioning on such post-treatment variables blocks mediated channels and can open collider paths, and this contamination is not confined to *γ*: once a collider is conditioned on, *no* coefficient in the equation retains a causal reading. We therefore define the estimand of Eq (3) precisely and modestly. Its coefficients are those of the linear projection—the best linear predictor—of adult symptoms on the conditioning set, a well-defined population object that answers a purely predictive question: does the Round-6 shock retain *incremental predictive power* once the persistence of symptoms and of material hardship is held fixed? No coefficient in Eq (3), including those on the added regressors, is given a causal or partial-effect interpretation. Because this predictive reading is deliberately weak, we complement it with a collider-free characterisation of the same channels: a chain of persistence regressions in which each link—adult food insecurity on pandemic food insecurity, and each adult symptom score on its own pandemic-period lag—conditions only on the predetermined vector and a single lagged variable (Appendix Table A8).

#### Latent-factor measurement model

To address measurement error in early human capital we estimate a confirmatory factor model in the spirit of the measurement system of Cunha et al. [5]. A latent early-health stock *θ_i_* is measured by three noisy indicators, the height-for-age *z*-scores of Rounds 1–3:

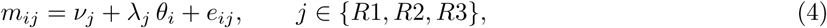

with the scale fixed by *λ_R_*_1_ = 1 and *e_ij_ ⊥ θ_i_*. The predicted factor score is then used in place of the single-round proxy in Eq (2). Because a single-factor model with three indicators is exactly identified—so that conventional fit indices are uninformative by construction—we also estimate an *overidentified* version that adds the Round-1 weight-for-age *z*-score as a fourth indicator, allowing the Round-1 height and weight uniquenesses to covary since they are measured at the same visit; this specification has one overidentifying degree of freedom and delivers testable fit statistics—the root mean squared error of approximation (RMSEA), the comparative fit index (CFI), the Tucker–Lewis index (TLI), and the standardised root mean squared residual (SRMR)—reported in Appendix Table A1. As a further check on measurement error that does not restrict the Round 1–3 dynamics at all, we also instrument Round-1 height-for-age with Round-1 weight-for-age—a classical errors-in-variables correction using only infancy measurements.

#### Self-productivity: dynamic panel models

Self-productivity of the health stock—a central feature of the skill-formation technology [4]—implies a positive autoregressive coefficient in

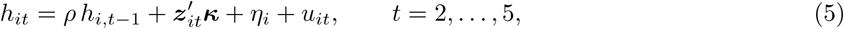

where *h_it_* is the height-for-age *z*-score in round *t*, *η_i_* a child fixed effect, and ***z****_it_* time-varying controls (the wealth index and round effects). Because the within estimator of *ρ* is biased in short panels [24], we report the difference generalised method of moments (GMM) estimator of Arellano and Bond [25], which instruments the first-differenced equation with lagged levels, exploiting the moment conditions

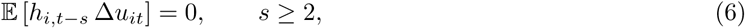

and the system-GMM estimator of Blundell and Bond [26], which augments these with lagged first-differences as instruments for the levels equation. Fixed- and random-effects estimates are compared with the Hausman test [27]; instrument validity is assessed with the Sargan test and serial correlation with the Arellano–Bond AR(2) statistic. Because a rejection of the AR(2) null implies that the *t −* 2 lags are themselves invalid instruments—the moment conditions in Eq (6) then fail for *s* = 2—we re-estimate the model restricting the instrument set to lags *t −* 3 and deeper, which remain valid under a first-order moving-average error structure such as classical anthropometric measurement error, and which also curbs instrument proliferation [28]. Both a full deep-lag set and a parsimonious single-lag set are reported (Appendix Table A4).

#### Heterogeneity across countries, cohorts, sexes, and the resilience hypothesis

To test proposition P3 and contextual heterogeneity we add a battery of interactions to Eq (2):

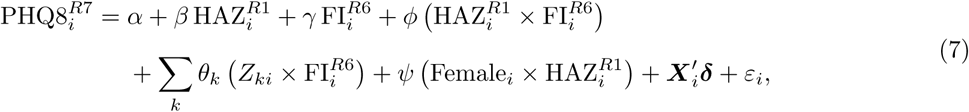

where *Z_k_* indexes sex and country. A negative and significant *ϕ* would support resilience (dynamic complementarity); the *θ_k_* capture differential gradients by group. In a parallel specification we interact the birth cohort indicator with both the shock and early human capital, which asks whether being hit by the pandemic at age 26 (Older Cohort) rather than at age 19 (Younger Cohort) changed the size of the gradient; note that age at the shock, age at measurement, and birth cohort are perfectly collinear in a two-cohort design, so any cohort contrast admits all three readings. Because Eq (7) does not interact height-for-age with country, and returns to early endowments may differ across countries, we treat the fully flexible check—re-estimating Eq (2) separately by country and by cohort—as the *primary* heterogeneity evidence, with the pooled interaction model as a complement. Given the number of contrasts examined (two outcomes, three countries, two cohorts, two sexes), all heterogeneity results are labelled exploratory, and the interaction *p* values are read against a Holm adjustment within this family.

#### Binary outcomes

Formal employment *F_i_*(holding a written contract) is modelled with a logit specification,

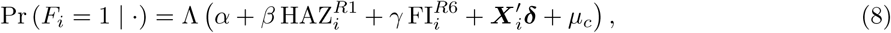

with Λ(*·*) the logistic cumulative distribution function; we report odds ratios and average marginal effects.

#### Mechanisms: mediation

Following the mechanism-decomposition logic of Heckman et al. [29] and the classical mediation framework [30], we decompose the early-capital association into a direct path and a path mediated by completed schooling *M_i_*:

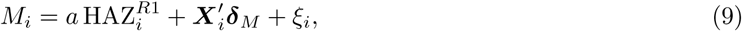

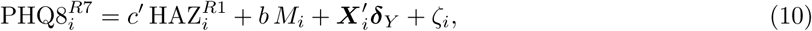

so that the indirect effect is *a b*, the direct effect is *c^′^*, and the total effect is *c^′^* + *a b*. Because the mediator and the outcome are both measured in Round 7, the errors *ξ_i_* and *ζ_i_* may be correlated through contemporaneous omitted variables (for example, a recent family shock affecting both school completion and mood), in which case *b* is not identified without further assumptions. We therefore accompany the baseline decomposition—which assumes sequential exogeneity, Cov(*ξ_i_, ζ_i_*) = 0—with a sensitivity analysis in the spirit of Imai et al. [31]: we fix the error correlation *ϱ* on a grid spanning *−*0.9 to 0.9—essentially the entire feasible range, so that the exercise is not conditioned on any assumption about the strength of contemporaneous confounding—re-estimate the system subject to Cov(*ξ_i_, ζ_i_*) = *ϱ σ_ξ_σ_ζ_*, and trace out the implied direct and indirect effects (Appendix Table A6). Two features make the exercise informative rather than merely illustrative. The first-stage coefficient *a* involves only the Round-1 endowment and Round-7 schooling and is invariant to *ϱ* by construction; and the indirect effect *a b* is the product of this invariant, precisely estimated near-zero quantity with the *ϱ*-sensitive *b*, so its magnitude is bounded close to zero for *any* admissible value of the confounding correlation.

#### Attrition and inverse-probability weighting

Round 7 was not fielded in Vietnam by design, but within Ethiopia, India, and Peru roughly one fifth of the Round-1 sample is not observed for adult mental health, and ordinary least squares is consistent only if drop-out is unrelated to the outcome equation errors. We therefore estimate a probit model of Round-7 retention on the full predetermined Round-1 vector, construct inverse-probability weights (IPW) as the reciprocal of the fitted retention probability (trimmed at the 99th percentile), and re-estimate the main gradient models by weighted least squares [32]. Selection on Round-1 observables alone is, however, a strong assumption over a 22-year window, because drop-out may respond to intermediate, time-varying shocks. For the estimation samples that condition on the Round 6 phone survey—precisely the samples in which the gradient is estimated—we therefore also construct *sequential* weights from a probit of Round-7 retention on the Round-1 vector augmented with the Round-6 outcomes themselves (pandemic-period PHQ-8, GAD-7, and food insecurity). Conditioning retention on the lagged outcome absorbs time-varying selection operating through the observed trajectory of mental health or hardship up to 2020—a sequential missing-at-random assumption substantially weaker than selection on childhood observables—while selection on post-2020 unobservables orthogonal to the lagged outcome remains untestable (Appendix Table A5). Because pandemic food insecurity itself predicts drop-out, weighting alone cannot settle the matter; we therefore also compute Lee-style trimming bounds for the binary moderate/severe exposure, which bound the mean contrast among always-retained participants under a monotonicity assumption and no selection model at all. Finally, since the weighted regressions treat the estimated weights as fixed, we re-estimate the entire two-step sequential procedure (retention probit plus weighted regression) by cluster bootstrap to incorporate first-stage estimation error into the standard errors.

#### Inference

Standard errors of all regression models are clustered at the sentinel-site level to account for within-community correlation [33]; the estimation samples contain 60 clusters in the baseline models and 56 clusters in the models conditioning on the Round 6 phone survey. One, two, and three asterisks denote statistical significance at the 10%, 5%, and 1% levels. Specification is assessed with Breusch–Pagan, Ramsey RESET, and link tests for linear models, and with the Hosmer–Lemeshow test and the area under the ROC curve for logit models; cross-country differences in means are tested with Wald tests on the cluster-robust variance matrix (one-way ANOVA would ignore the clustered design), and all diagnostic statistics are collected in Appendix Table A1. Because the country-specific samples contain only 16–20 clusters—a range in which conventional cluster-robust inference over-rejects—every country-specific food-insecurity coefficient and the pooled country-interaction test are additionally evaluated with the wild cluster bootstrap-*t* (Webb six-point weights, 999 replications, null imposed). Missing data are handled by listwise deletion within each model, so estimation samples vary across tables; adult height is additionally subjected to adult plausibility limits (120–210 cm). Item-level PHQ-8 and GAD-7 responses are not included in deposit SN 9543, which contains only the four constructed country files; the item data reside in the separate round-specific Young Lives deposits, outside the scope of this replication package, so formal measurement-invariance testing across countries and languages is not possible here; as a minimum comparability check we re-express the country-specific gradients with outcome and exposure standardised within country, and we flag that differential item functioning and floor effects cannot be ruled out. The Breusch–Pagan test strongly rejects homoskedasticity in every linear model, which motivates the cluster-robust inference used throughout; the RESET and link tests point to mild non-linearity of the conditional mean, as expected for bounded symptom counts, so we verify that the key associations are robust to the binary coding of the shock (Table 4, column 3). The mediation system is estimated by maximum likelihood. All analyses use Stata 17.

## Results

### Descriptive comparisons across countries and cohorts

Table 2 compares the four study countries on childhood conditions, pandemic exposure, and adult outcomes; cluster-robust Wald tests reject equality of means across countries for every row except mean early height-for-age itself (*p* = 0.32), whose apparent country differences do not survive the clustered design. Three contrasts organise the comparison. First, initial conditions: because the height-for-age means themselves are statistically indistinguishable across countries under the clustered design, the initial-conditions contrast rests on the indicators that do survive it—early stunting was most prevalent in Ethiopia (39%) and least in Vietnam (23%; *p* = 0.005), and maternal schooling was far higher in Peru (7.0 grades) than in Ethiopia or India (about 2.7–2.8). Second, pandemic exposure: food insecurity during COVID-19 was most widespread in Ethiopia (mean 2.06 on the 1–4 scale; 19% moderate or severe) against roughly 5% moderate or severe in India and Peru—and Ethiopian households remain the most food-insecure in adulthood (2.17 at Round 7). Third, adult outcomes: mean adult PHQ-8 is highest in Peru (3.39), intermediate in Ethiopia (2.11), and lowest in India (1.55), with the same ordering for GAD-7 (3.96, 2.82, 1.73); formal employment ranges from 25% of workers in Peru and 16% in Ethiopia to 7% in India. Cohort patterns also differ by country: in Ethiopia and India the older cohort reports slightly more symptoms than the younger (for example, PHQ-8 of 2.23 versus 2.06 in Ethiopia), whereas in Peru the younger cohort reports more (3.55 versus 2.87). Women report substantially worse mental health than men (PHQ-8 of 2.67 versus 1.95; GAD-7 of 3.25 versus 2.31).

**Table 2.**
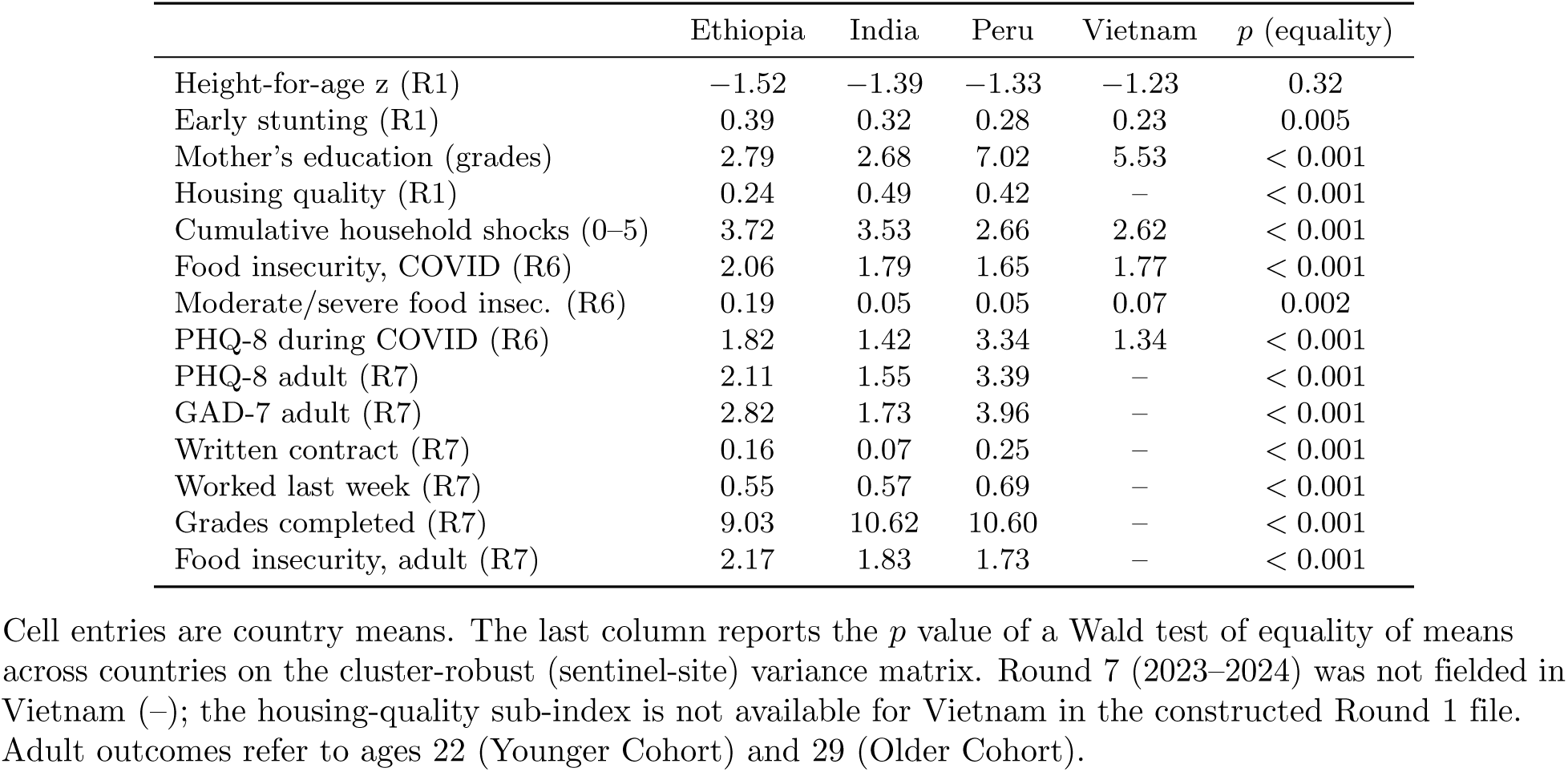
Country profiles: means of childhood conditions, pandemic exposure, and adult outcomes.

|  | Ethiopia | India | Peru | Vietnam | *p* (equality) |
| --- | --- | --- | --- | --- | --- |
| Height-for-age *z* (R1) | −1.52 | −1.39 | −1.33 | −1.23 | 0.32 |
| Early stunting (R1) | 0.39 | 0.32 | 0.28 | 0.23 | 0.005 |
| Mother’s education (grades) | 2.79 | 2.68 | 7.02 | 5.53 | < 0.001 |
| Housing quality (R1) | 0.24 | 0.49 | 0.42 | – | < 0.001 |
| Cumulative household shocks (0–5) | 3.72 | 3.53 | 2.66 | 2.62 | < 0.001 |
| Food insecurity, COVID (R6) | 2.06 | 1.79 | 1.65 | 1.77 | < 0.001 |
| Moderate/severe food insecurity (R6) | 0.19 | 0.05 | 0.05 | 0.07 | 0.002 |
| PHQ-8 during COVID (R6) | 1.82 | 1.42 | 3.34 | 1.34 | < 0.001 |
| PHQ-8 adult (R7) | 2.11 | 1.55 | 3.39 | – | < 0.001 |
| GAD-7 adult (R7) | 2.82 | 1.73 | 3.96 | – | < 0.001 |
| Written contract (R7) | 0.16 | 0.07 | 0.25 | – | < 0.001 |
| Worked last week (R7) | 0.55 | 0.57 | 0.69 | – | < 0.001 |
| Grades completed (R7) | 9.03 | 10.62 | 10.60 | – | < 0.001 |
| Food insecurity, adult (R7) | 2.17 | 1.83 | 1.73 | – | < 0.001 |

Cell entries are country means. The last column reports the *p* value of a Wald test of equality of means across countries on the cluster-robust (sentinel-site) variance matrix. Round 7 (2023–2024) was not fielded in Vietnam (–); the housing-quality sub-index is not available for Vietnam in the constructed Round 1 file. Adult outcomes refer to ages 22 (Younger Cohort) and 29 (Older Cohort).

### The long reach of early human capital

Early height-for-age is a powerful predictor of adult *stature*. In the spline specification of Table 3 (with adult height subjected to adult plausibility limits) the away-from-the-knot slope is 1.20 cm per standard deviation (*p <* 0.01); with the continuous score alone—the total association, which is the policy-relevant quantity—it is 1.60 cm per standard deviation (standard error 0.11). Early height-for-age is *not*, however, associated with adult formal employment: with the continuous score alone the odds ratio is 1.05 (*p* = 0.19; Table 7, discussed below), and the apparent stunting odds ratio in the spline form is a segment of the same spline rather than a separate construct—maternal schooling (odds ratio 1.06, *p <* 0.01) is the robust early predictor. Nor is early height-for-age associated with adult depressive symptoms (Table 4, column 1) or anxiety (Table 5, column 1).

**Table 3.** Validation: early human capital predicts adult height but not schooling.

|  | (1)<br>Grades completed (R7) | (2)<br>Adult height, cm (R7) |
| --- | --- | --- |
| Height-for-age z (R1) | 0.031<br>(0.033) | 1.202***<br>(0.137) |
| Early stunting (R1) | −0.048<br>(0.098) | −1.697***<br>(0.211) |
| Housing quality (R1) | 0.611**<br>(0.278) | 1.190**<br>(0.457) |
| Services access (R1) | 0.416*<br>(0.247) | −0.623<br>(0.456) |
| Consumer durables (R1) | 0.302<br>(0.294) | 2.280***<br>(0.569) |
| Mother’s education (grades) | 0.035***<br>(0.010) | 0.046**<br>(0.022) |
| Father’s education (grades) | 0.057***<br>(0.011) | 0.039<br>(0.024) |
| Female | 0.008<br>(0.098) | −12.795***<br>(0.212) |
| Urban | 0.154<br>(0.173) | −0.082<br>(0.368) |
| Cumulative household shocks (0–5) | −0.011<br>(0.039) | −0.032<br>(0.092) |
| Younger cohort | 0.376***<br>(0.091) | 0.151<br>(0.217) |
| Constant | 8.206***<br>(0.231) | 173.468***<br>(0.537) |
| Country fixed effects | Yes | Yes |
| Observations | 6019 | 5835 |
| R-squared | 0.182 | 0.615 |
| Adjusted R-squared | 0.180 | 0.614 |

Cluster-robust standard errors in parentheses (60 sentinel-site clusters). Adult height is subjected to adult plausibility limits (120–210 cm; one record removed). Because early stunting is the indicator $1[\text{HAZ} < -2]$ , the two anthropometric regressors form a linear spline with a knot at $-2$ ; the height-for-age coefficient is the slope away from the knot, and the total association with the continuous score alone is 1.60 cm per standard deviation (standard error 0.11). \* $p < 0.10$ , \*\* $p < 0.05$ , \*\*\* $p < 0.01$ .

**Table 4.**
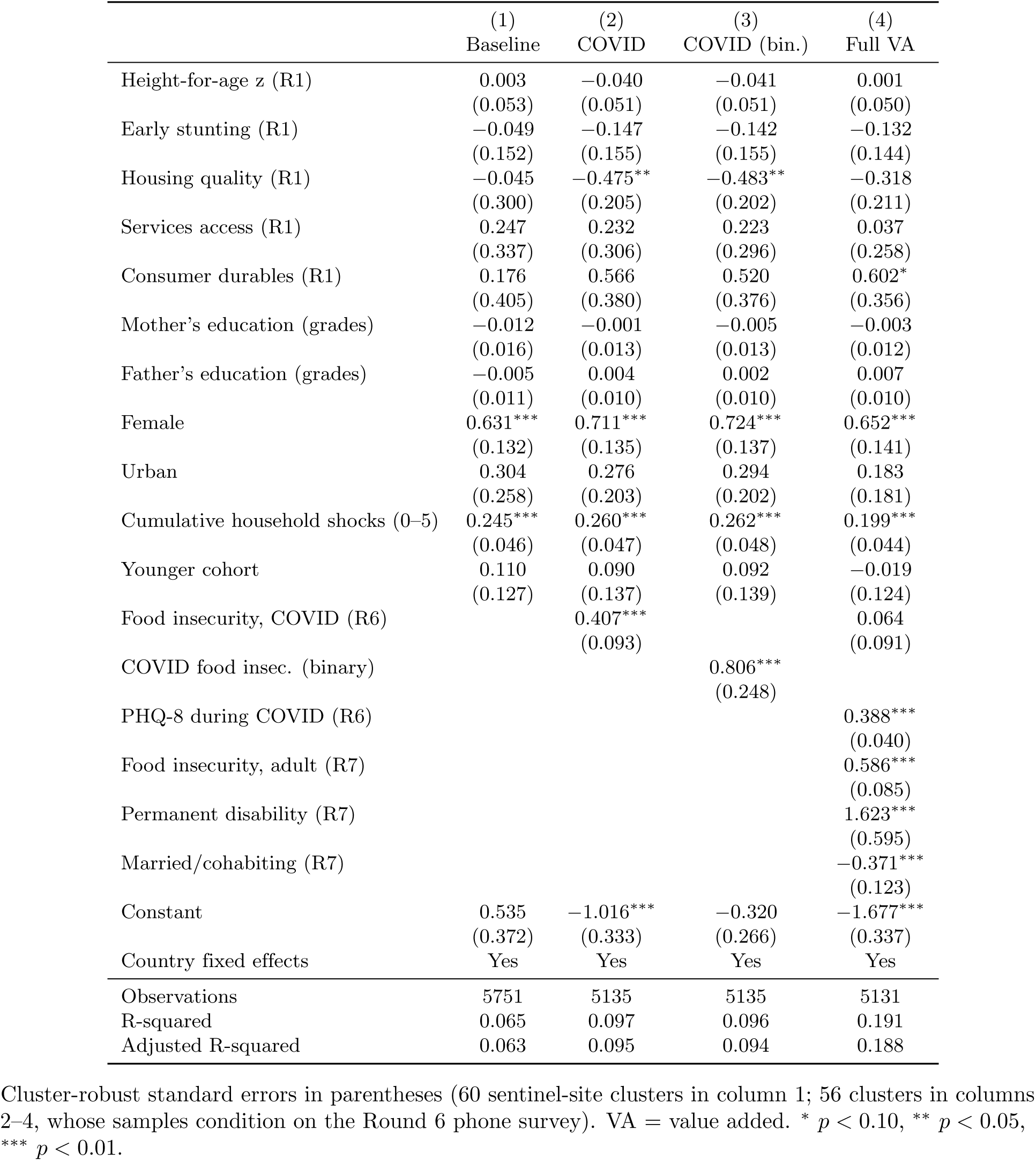
Early human capital, the COVID-19 shock, and adult depression (PHQ-8).

**Table 5.** Early human capital, the COVID-19 shock, and adult anxiety (GAD-7).

|  | (1)<br>Baseline | (2)<br>COVID | (3)<br>Full VA |
| --- | --- | --- | --- |
| Height-for-age z (R1) | 0.027<br>(0.060) | −0.019<br>(0.047) | 0.019<br>(0.047) |
| Early stunting (R1) | 0.082<br>(0.150) | −0.097<br>(0.143) | −0.091<br>(0.140) |
| Housing quality (R1) | 0.364<br>(0.398) | −0.173<br>(0.254) | −0.055<br>(0.222) |
| Services access (R1) | 0.100<br>(0.443) | 0.017<br>(0.287) | −0.149<br>(0.222) |
| Consumer durables (R1) | −0.256<br>(0.510) | 0.474<br>(0.375) | 0.658**<br>(0.316) |
| Mother's education (grades) | −0.035**<br>(0.014) | −0.014<br>(0.013) | −0.019<br>(0.012) |
| Father's education (grades) | −0.020<br>(0.016) | −0.008<br>(0.010) | −0.006<br>(0.010) |
| Female | 0.837***<br>(0.143) | 0.935***<br>(0.152) | 0.774***<br>(0.144) |
| Urban | 0.680*<br>(0.364) | 0.639***<br>(0.179) | 0.496***<br>(0.143) |
| Cumulative household shocks (0–5) | 0.207**<br>(0.080) | 0.290***<br>(0.045) | 0.208***<br>(0.040) |
| Younger cohort | −0.238*<br>(0.122) | −0.195<br>(0.126) | −0.121<br>(0.132) |
| Food insecurity, COVID (R6) |  | 0.414***<br>(0.095) | −0.002<br>(0.088) |
| GAD-7 during COVID (R6) |  |  | 0.427***<br>(0.031) |
| Food insecurity, adult (R7) |  |  | 0.538***<br>(0.088) |
| Permanent disability (R7) |  |  | 0.980**<br>(0.426) |
| Married/cohabiting (R7) |  |  | −0.238**<br>(0.113) |
| Constant | 1.523**<br>(0.707) | −0.975***<br>(0.325) | −1.512***<br>(0.330) |
| Country fixed effects | Yes | Yes | Yes |
| Observations | 5885 | 5132 | 5128 |
| R-squared | 0.082 | 0.134 | 0.238 |
| Adjusted R-squared | 0.080 | 0.132 | 0.235 |

Cluster-robust standard errors in parentheses (60 sentinel-site clusters in column 1; 56 clusters in columns 2–3, whose samples condition on the Round 6 phone survey). VA = value added. \* $p < 0.10$ , \*\* $p < 0.05$ , \*\*\* $p < 0.01$ .

We stress that this is a *precise* null, not an absence of evidence: the baseline coefficient of 0.003 (standard error 0.053) confines the association to within *±*0.03 standard deviations of PHQ-8 per standard deviation of infant height-for-age at the 95% level, and the conclusion is unchanged with the continuous score alone (0.015, standard error 0.035), with the latent-factor scores (see Mechanisms), and when Round-1 height-for-age is instrumented with Round-1 weight-for-age to purge classical measurement error (0.031, standard error 0.058). Adult mental health appears to be governed by more proximate circumstances than by early nutritional endowments.

### The medium-term gradient of pandemic food insecurity

Household food insecurity during the pandemic is robustly associated with higher adult depression three to four years later: each step on the four-point insecurity scale raises PHQ-8 by 0.41 points, or 0.12 standard deviations (95% CI 0.22–0.59, *p <* 0.01; Table 4, column 2), and the binary moderate/severe indicator raises it by 0.81 points (0.23 standard deviations; column 3). The same pattern holds for anxiety (Table 5, column 2: *β* = 0.41 points, 0.11 standard deviations, 95% CI 0.22–0.60, *p <* 0.01). The bounded-count and clinical-threshold models of Appendix Table A9 translate these magnitudes: the Poisson incidence-rate ratios are 1.20 (PHQ-8) and 1.18 (GAD-7) per step, and each step raises the probability of clinically relevant anxiety (GAD-7 *≥* 10; base rate 7.6%) by 1.9 percentage points (*p* = 0.001) and of clinically relevant depression (PHQ-8 *≥* 10; base rate 5.6%) by 1.0 percentage point (*p* = 0.07). The linear coding of the ordinal exposure is supported by the data: category indicators show a monotone dose–response over the well-populated range (0.30 points for “sometimes” and 1.08 for “often” short of enough food, relative to always eating enough; both *p <* 0.05; the most extreme category is rare and imprecisely estimated), and the Wald test does not reject the equidistance restriction (*p* = 0.19 for PHQ-8 and *p* = 0.12 for GAD-7; Appendix Table A7). Better childhood housing quality is protective in the pandemic-exposed sample (*−*0.48, *p <* 0.05, column 2); its near-zero coefficient in column 1 reflects sample composition rather than conditioning, since re-estimating the baseline on the column-2 sample yields *−*0.53 (standard error 0.20). In the value-added models (Table 4, column 4; Table 5, column 3) the pandemic-period coefficient attenuates to non-significance once lagged pandemic distress (*ρ* = 0.39 for PHQ-8 and 0.43 for GAD-7, both *p <* 0.01) and *continued* adult food insecurity (*β* = 0.59 and 0.54, both *p <* 0.01) are included, indicating that the association travels through the persistence of distress and of material hardship rather than as an independent “memory” of 2020; as set out in Methods, these value-added columns are linear-projection (predictive) decompositions—their added regressors are post-treatment mediators, and no coefficient in them carries a causal reading—so the magnitude of the gradient should be read from column 2. The collider-free persistence chain of Appendix Table A8 tells the same story with clean conditioning: pandemic food insecurity strongly predicts *adult* food insecurity (0.19, *p <* 0.01), and each pandemic-period symptom score strongly predicts its adult counterpart (0.41 for PHQ-8 and 0.47 for GAD-7, both *p <* 0.01), with each link conditioning only on the predetermined vector and a single lag. Disability (depression *β* = 1.62, *p <* 0.01), being unmarried, female sex, and cumulative childhood adversity (*β* = 0.20, *p <* 0.01) are all independent correlates of poor adult mental health.

How much of the gradient could a stable individual propensity to report both hardship and distress explain? The Round-6 data bound the answer. Conditional on the predetermined vector, the *contemporaneous* association between pandemic food insecurity and pandemic distress is 0.57 points of PHQ-8 and 0.72 points of GAD-7 per step. If that contemporaneous association reflected nothing but a stable trait that also drives adult symptoms, the implied medium-term gradient would be the contemporaneous association times the persistence of symptoms—0.57 *×* 0.41 *≈* 0.24 for PHQ-8 and 0.72 *×* 0.47 *≈* 0.34 for GAD-7—which falls short of the estimated 0.41 in both cases. This is an *illustrative benchmark*, not an upper bound: both inputs are attenuated by measurement error in the Round-6 score—the persistence coefficient understates the persistence of any stable component, and the contemporaneous association understates its link to the exposure—so the trait channel could be larger than the benchmark suggests. What the calculation establishes is more modest but still useful: the observed gradient is not *numerically* the product of the observed 2020 cross-section and the observed persistence, so some part of it operates outside that simple chain. What remains is bounded rather than identified. Because Round-6 distress is measured *simultaneously* with the exposure, conditioning on it in the value-added column plays a dual role—it blocks a genuine mediator (persistence of distress) *and* controls the reporting-propensity confounder—so, *provided that conditioning on Round-6 distress does not itself open a collider path*, the two specifications can be read as a bracket: the value-added coefficient (0.06, n.s.) and the unconditional gradient (0.41) bound the causal parameter under the polar readings of Round-6 distress as pure mediator or pure confounder.

The gradient is also robust to non-random attrition. A probit of Round-7 retention on the Round-1 covariates shows that follow-up is predicted by cumulative household shocks, cohort, sex, and country, but *not* by early height-for-age, stunting, or parental education; re-estimating the column-2 models with the implied inverse-probability weights moves the food-insecurity coefficient from 0.41 to 0.38 for PHQ-8 and from 0.41 to 0.37 for GAD-7 (both *p <* 0.01; Appendix Table A5), leaving every conclusion intact. The sequential weights—which additionally condition Round-7 retention on the Round-6 outcomes themselves—sharpen this conclusion: pandemic-period depression and anxiety do *not* predict subsequent drop-out (both *p >* 0.5), pandemic food insecurity does (*−*0.27, *p <* 0.01), and the sequentially reweighted gradients are 0.40 for both outcomes, indistinguishable from the unweighted estimates. Because the exposure itself predicts drop-out, we additionally compute Lee-style trimming bounds for the binary exposure, which require no selection model: with retention of 92.9% among exposed and 98.0% among unexposed households (trimming fraction 0.052), the bounds on the mean PHQ-8 contrast among always-retained participants are [0.65, 1.17]—both ends positive, the lower end equal to the untrimmed difference—so differential attrition cannot explain the association away. These bounds are *unadjusted* mean contrasts by construction, so they are not directly comparable to the covariate-adjusted binary coefficient of 0.81 in Table 4 and should not be read as an attenuated version of it. A cluster bootstrap of the full two-step sequential procedure (retention probit plus weighted regression; 499 replications) confirms that treating the estimated weights as fixed does not understate the uncertainty: the bootstrap standard error is 0.090 against the analytic 0.093, with a percentile 95% CI of [0.23, 0.59].

### Where and for whom: countries, cohorts, sex, and resilience

All results in this subsection are exploratory, as pre-announced in Methods. The pooled interaction model (Table 6) shows that the resilience term—early human capital *×* pandemic food insecurity—is positive but not statistically significant (*ϕ* = 0.070, *p* = 0.28), so the dynamic-complementarity hypothesis (P3) is *not* supported; nor does the height-for-age association with depression differ by sex (*p* = 0.63), nor the food-insecurity slope (female *×* food insecurity *p* = 0.83; women sit about 0.7 points above men at every exposure level, Fig 2, panel B).

**Fig 2.**
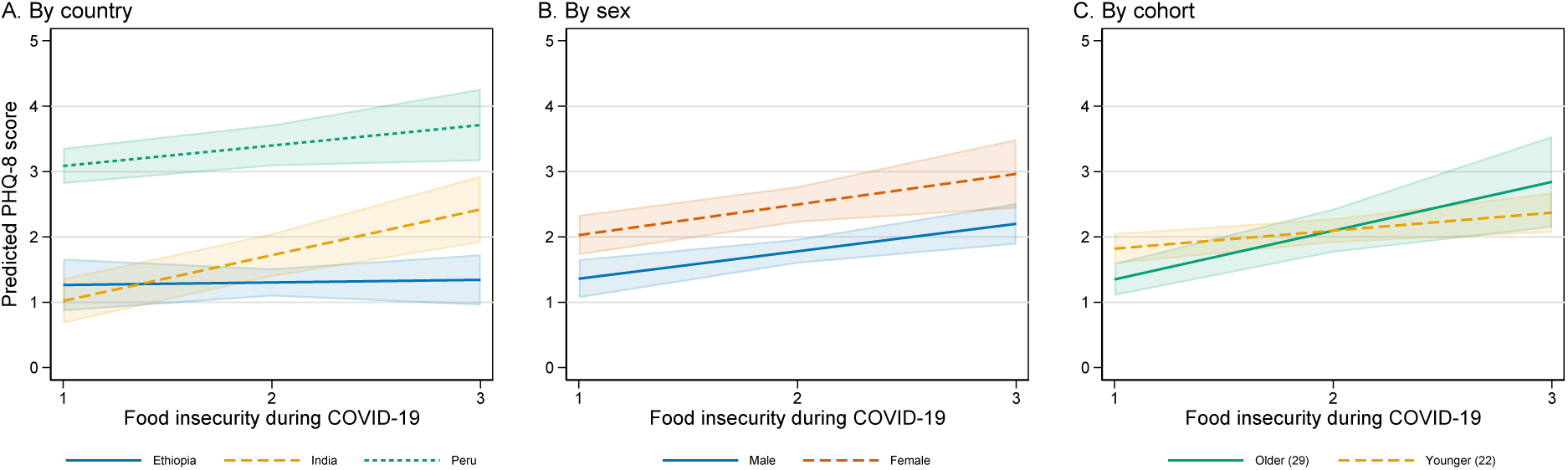
Predicted adult depression by pandemic food insecurity, across countries, by sex, and by cohort. Predictive margins with 95% confidence bands, shown over the well-supported range of the exposure (categories 1–3; the most severe category holds under 2% of the sample and is omitted). A: by country (from the interaction model of Table 6)—the gradient is flat in Ethiopia, steepest in India, and intermediate in Peru, which has the highest overall symptom level. B: by sex—women report more symptoms at every exposure level, but the slopes do not differ. C: by cohort (from the cohort-interaction model of Appendix Table A3)—the gradient appears steeper for the Older Cohort. Country and cohort contrasts are exploratory (see text).

**Table 6.** Heterogeneity of the COVID-19 mental-health gradient (PHQ-8).

|  | (1) |
| --- | --- |
| Height-for-age z (R1) | −0.156<br>(0.131) |
| Food insecurity, COVID (R6) | 0.112<br>(0.213) |
| HAZ × Food insecurity (R6) | 0.070<br>(0.064) |
| Female × Food insecurity (R6) | 0.048<br>(0.222) |
| India × Food insecurity (R6) | 0.661***<br>(0.217) |
| Peru × Food insecurity (R6) | 0.274<br>(0.220) |
| Female × HAZ | −0.025<br>(0.052) |
| Female | 0.585<br>(0.406) |
| Housing quality (R1) | −0.451**<br>(0.208) |
| Cumulative shocks | 0.258***<br>(0.047) |
| Observations | 5135 |
| Adjusted R-squared | 0.096 |

The primary heterogeneity evidence is the country-by-country estimation of Eq (2) (Fig 3; full models in Appendix Table A2). The gradient is essentially zero in Ethiopia (0.00, 95% CI *−*0.31 to 0.31), largest in India (0.58, 95% CI 0.29–0.87), and intermediate in Peru (0.41, 95% CI 0.10–0.72)—even though India shows the *lowest* average symptom levels—and the anxiety outcome replicates the ordering exactly (*−*0.04, 0.60, and 0.42). Because these subsamples contain only 16–20 clusters, we re-evaluate every coefficient with the wild cluster bootstrap-*t*: the pattern survives, with *p*_wild_ = 0.001 (PHQ-8) and *<* 0.001 (GAD-7) in India, *p*_wild_ = 0.013 and 0.033 in Peru, and *p*_wild_ *>* 0.75 in Ethiopia; the pooled country-interaction test degrades only mildly, from *p* = 0.012 to *p*_wild_ = 0.019. The ordering also survives the minimum

**Fig 3.**
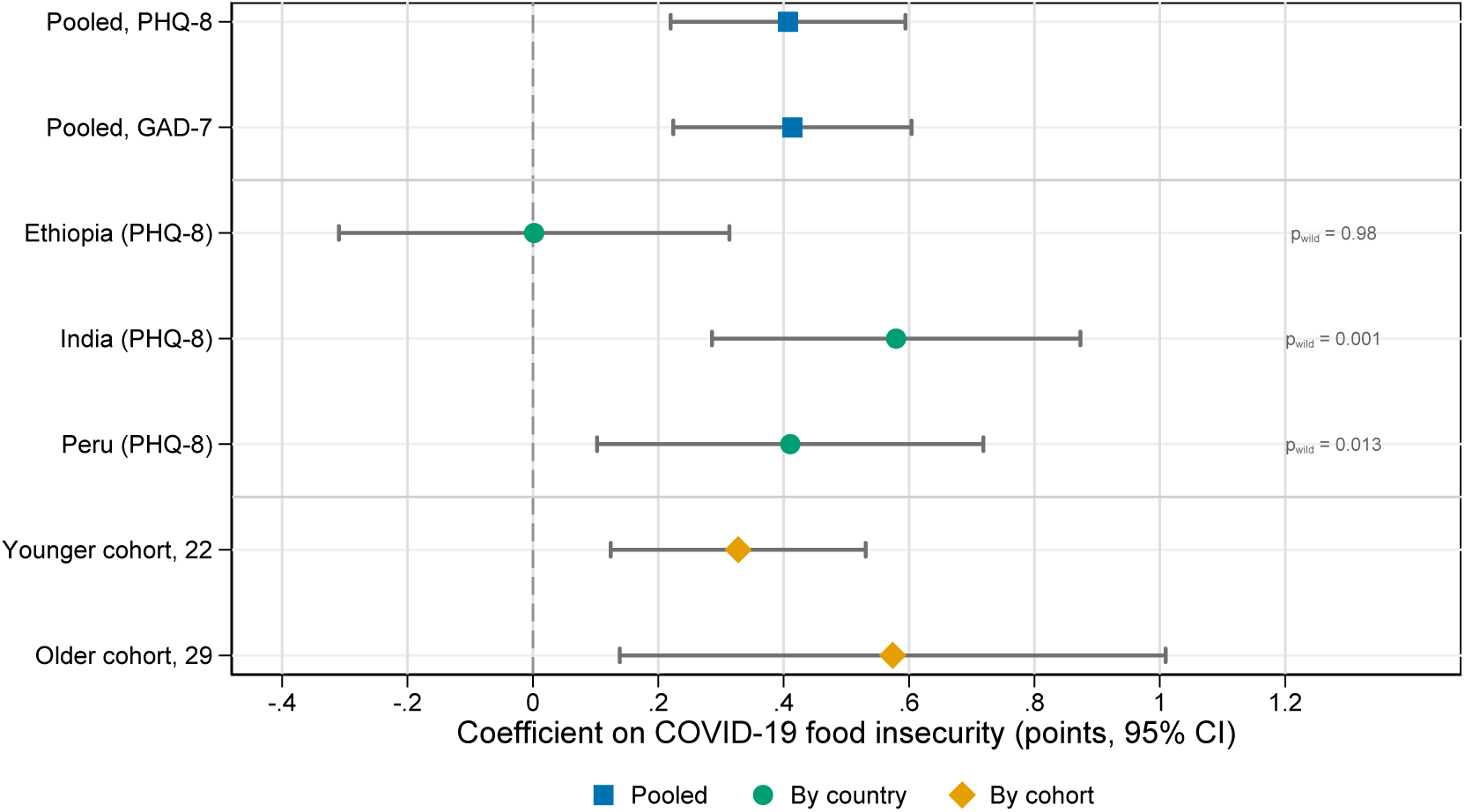
The COVID-19 food-insecurity gradient, pooled and by country and cohort. Coefficients on Round-6 food insecurity (1–4 scale) from Eq (2) estimated on the pooled sample (PHQ-8 and GAD-7) and separately by country and by birth cohort (PHQ-8), with 95% confidence intervals based on sentinel-site-clustered standard errors; the annotations report wild cluster bootstrap-*t p* values (Webb weights, 999 replications) for the country subsamples, whose 16–20 clusters make conventional cluster-robust inference unreliable. Subsample models omit the fixed effects of the dimension they split on.

measurement-comparability check: with outcome and exposure standardised within country, the gradients are 0.000 (Ethiopia), 0.120 (India), and 0.061 (Peru) standard deviations per within-country standard deviation of exposure for PHQ-8, and *−*0.009, 0.123, and 0.058 for GAD-7. What no re-scaling can rule out, however, is differential item functioning across languages and floor effects—India combines the lowest symptom level with the steepest slope, exactly the signature such artefacts would produce—so we read the country contrast as a robust *statistical* pattern whose substantive interpretation remains open. Fig 2 (panel A) plots the implied predicted depression profiles by country over the well-supported exposure range.

The two birth cohorts were hit by the pandemic at very different life stages—roughly age 19 (Younger Cohort) versus age 26 (Older Cohort)—although, as noted in Methods, age at the shock, age at measurement, and birth cohort are perfectly collinear, so what follows admits all three readings. Interacting cohort with the shock, the food-insecurity gradient on depression is 0.75 points per step (*p <* 0.01) for the Older Cohort but 0.47 points *smaller* for the Younger Cohort (interaction *p* = 0.038); for anxiety the corresponding figures are 0.79 and *−*0.53 (interaction *p* = 0.013). The cohort-specific estimates are more modest and visibly overlap: 0.57 (95% CI 0.14–1.01) for the Older Cohort against 0.33 (95% CI 0.12–0.53) for the Younger Cohort for depression, and 0.72 against 0.29 for anxiety (Fig 3; predicted profiles in Fig 2, panel C; full models in Appendix Table A3). The gap between the interaction estimate (0.75) and the subsample estimate (0.57) for the same group signals numerical fragility, the interaction *p* values are marginal, and within the family of heterogeneity contrasts examined here a Holm adjustment leaves none of them significant at conventional levels. We therefore state the cohort result deliberately weakly: the gradient *appears larger* in the cohort hit as young adults—already out of school and entering labour and marriage markets—but the intervals overlap and the contrast is exploratory. In contrast, the (null) association of early height-for-age with adult mental health is the same in both cohorts (interaction *p* = 0.89 for PHQ-8 and *p* = 0.26 for GAD-7).

Fig 4 completes the picture for the resilience hypothesis: the marginal effect of pandemic food insecurity on adult depression is, if anything, *increasing* in early height-for-age (from 0.33 at HAZ = *−*3 to 0.68 at HAZ = 2), the opposite of the protective moderation that dynamic complementarity would imply, though the slope is imprecise (*p* = 0.28). Read as an equivalence statement rather than a non-rejection, the interaction estimate (*ϕ* = 0.070, standard error 0.064; 95% CI *−*0.055 to 0.195) rules out any buffering stronger than about *−*0.06 points of gradient per unit of early height-for-age: even the most favourable end of the interval would offset only a seventh of the pooled gradient per standard deviation of endowment.

**Fig 4.**
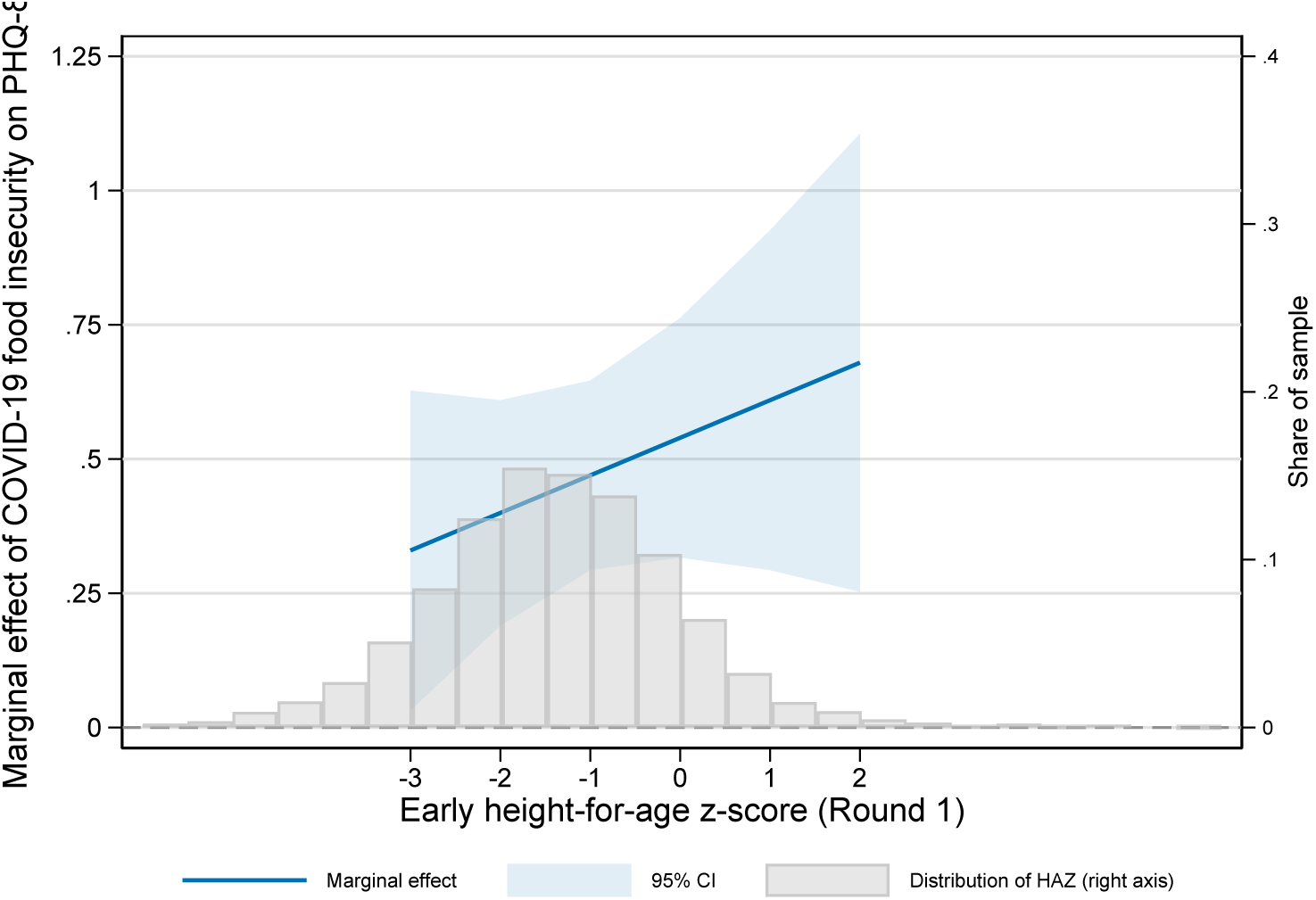
No evidence that early human capital buffers the pandemic shock. Marginal effect of Round-6 food insecurity on adult PHQ-8 at different levels of early height-for-age, from the interaction model of Table 6, with a 95% confidence band; the grey histogram (right axis) shows the distribution of early height-for-age in the estimation sample, so the profile can be read against its empirical support. A buffering (resilience) effect would imply a downward-sloping profile; the estimated profile is mildly upward-sloping and never distinguishable from a constant effect.

### Formal employment and the persistence of the health stock

Maternal schooling is the robust early predictor of adult formal employment (Table 7); the logit models show acceptable fit (area under the ROC curve 0.72; Hosmer–Lemeshow *p* = 0.08). The apparent contrast between the null height-for-age coefficient and the significant stunting odds ratio should not be read as two constructs behaving differently: because stunting is the indicator 1[HAZ *< −*2], the pair forms a spline in early height-for-age, and the stunting odds ratio is a segment of that spline rather than a separate finding. Two conditioning caveats apply: the outcome is defined only among participants who work, and the estimation sample is further reduced by covariate missingness (from 4,442 workers with contract information in Table 1 to 3,716 with full covariates), so selection into employment is not modelled and these are conditional-on-working associations. Pandemic food insecurity is unrelated to holding a written contract three to four years later (odds ratio 0.98, n.s.), suggesting the association is specific to mental health rather than operating through formal labour-market attachment. Members of the Younger Cohort, observed at age 22, are much less likely to hold a written contract than the Older Cohort at 29 (odds ratio 0.47, *p <* 0.01), consistent with gradual formalisation over the life course.

**Table 7.** Early human capital and adult formal employment (odds ratios).

|  | (1)<br>Baseline | (2)<br>COVID |
| --- | --- | --- |
| Height-for-age z (R1) | 0.982<br>(0.044) | 1.002<br>(0.051) |
| Early stunting (R1) | 0.752**<br>(0.092) | 0.753**<br>(0.101) |
| Housing quality (R1) | 1.219<br>(0.350) | 1.600*<br>(0.453) |
| Services access (R1) | 1.005<br>(0.285) | 0.968<br>(0.316) |
| Consumer durables (R1) | 1.339<br>(0.414) | 1.341<br>(0.391) |
| Mother's education (grades) | 1.063***<br>(0.015) | 1.063***<br>(0.018) |
| Father's education (grades) | 1.006<br>(0.015) | 1.004<br>(0.016) |
| Female | 1.155<br>(0.146) | 1.146<br>(0.147) |
| Urban | 1.200<br>(0.233) | 1.025<br>(0.208) |
| Cumulative household shocks (0–5) | 0.983<br>(0.046) | 0.998<br>(0.051) |
| Younger cohort | 0.468***<br>(0.062) | 0.466***<br>(0.062) |
| Food insecurity, COVID (R6) |  | 0.982<br>(0.093) |
| Country fixed effects | Yes | Yes |
| Observations | 3716 | 3256 |
| Pseudo R-squared | 0.103 | 0.108 |

The dynamic panel models are summarised in Table 8. In the baseline estimators the lagged height-for-age coefficient is nearly identical (*ρ* = 0.20, *p <* 0.01, system-GMM; *ρ* = 0.19, *p <* 0.01, difference-GMM), the wealth index enters at about 0.40, and the Hausman test decisively favours fixed effects (*χ*^2^(5) = 390.9, *p <* 0.01), confirming that unobserved child heterogeneity is correlated with household wealth. The baseline instruments, however, fail their own validity tests: the Arellano–Bond AR(2) statistic rejects the null of no second-order autocorrelation (*p* = 0.047)—which mechanically invalidates the moment conditions of Eq (6) at *s* = 2—and the Sargan test rejects the overidentifying restrictions (*χ*^2^(8) = 168.8, *p <* 0.01). Following the strategy set out in Methods, Appendix Table A4 re-estimates the model with the instrument set restricted to lags *t −* 3 and deeper. The AR(2) test then no longer rejects (*p* = 0.17 with the full deep-lag set; *p* = 0.22 with a single lag-3 instrument), as instrument validity requires, but the persistence estimate collapses to essentially zero (*ρ* = *−*0.02 and *−*0.01, standard errors 0.05) while the wealth coefficient strengthens to 0.45, and the Sargan test continues to reject. The implication must be stated without euphemism: a rejection of the overidentifying restrictions means the instruments are correlated with the structural error, so the GMM estimator is inconsistent and its point estimates—the baseline *ρ* = 0.19 *and* the near-zero deep-lag *ρ* alike—carry no interpretable content, descriptive or otherwise. The value of Appendix Table A4 is accordingly not any coefficient but the specification cascade itself: the moment structure of a stationary AR(1) in height-for-age at the round frequency—with rounds three to four years apart at very different ages, and anthropometry measured with error—is rejected by these data in every instrument configuration tried. We therefore draw no conclusion about round-to-round self-productivity in either direction. The long-reach evidence of Table 3, which compares levels 21 years apart and involves no internal instruments, is unaffected by this caveat.

**Table 8.** Persistence of the health stock: static and dynamic panel models.

|  | (1)<br>FE | (2)<br>RE | (3)<br>AB-GMM | (4)<br>SYS-GMM |
| --- | --- | --- | --- | --- |
| Wealth index | 0.281***<br>(0.052) | 0.928***<br>(0.040) | 0.398***<br>(0.051) | 0.400***<br>(0.052) |
| Lagged height-for-age z |  |  | 0.191***<br>(0.013) | 0.200***<br>(0.009) |
| Observations | 28567 | 28567 | 16527 | 22404 |

Height-for-age z-score, Younger Cohort, Rounds 1–5; round effects included. FE = fixed effects; RE = random effects; AB-GMM = Arellano–Bond difference-GMM; SYS-GMM = Blundell–Bond system-GMM. FE and RE use conventional standard errors (as required by the Hausman test); AB-GMM uses robust standard errors clustered by child. \* $p < 0.10$ , \*\* $p < 0.05$ , \*\*\* $p < 0.01$ .

### Mechanisms: latent factor and mediation

The confirmatory factor model of Eq (4) yields strong standardised loadings of the early health stock on the Round 1–3 height-for-age *z*-scores (0.67, 0.90, and 0.84; all *p <* 0.01); because the three-indicator model is just-identified, conventional fit indices are uninformative by construction. The overidentified four-indicator version—adding Round-1 weight-for-age, with the two Round-1 uniquenesses allowed to covary—fits the data well by conventional standards (RMSEA 0.044, probability RMSEA *≤* 0.05 of 0.70; CFI 0.999; TLI 0.994; SRMR 0.005; standardised loadings 0.67, 0.89, 0.84, and 0.54; Appendix Table A1). The factor score from either model is, like the single-round proxy, unrelated to adult depression (three-indicator *β*^^^ = 0.01; four-indicator *β*^^^ = 0.000, *p* = 0.99), so attenuation bias from measurement error does not explain the null long reach into mental health. The mediation system of Eqs (9)–(10) (Table 9) shows that completed schooling is protective against adult depression (*b* = *−*0.069, *p <* 0.01), but because early height-for-age does not raise completed schooling conditional on the covariates (*a* = 0.027, n.s.), the indirect path is small and non-significant (indirect effect *≈ −*0.002). Because the mediator and the outcome are both measured at Round 7, this decomposition assumes sequential exogeneity—and the mediator is itself censored from above (mean 10.1 grades against a maximum of 12), which attenuates *a*; completed schooling was in any case the only feasible mediator, since earnings are not included in the constructed files. The sensitivity analysis described in Methods quantifies what error correlation would do to the decomposition (Appendix Table A6): as *ϱ* sweeps essentially the entire feasible range, from *−*0.9 to 0.9, the path *b* swings from +1.33 to *−*1.47—confirming that *b* is simply not identified under simultaneity—yet *a* is invariant at 0.027 by construction (it links the Round-1 endowment to Round-7 schooling and does not involve the outcome equation), so the implied indirect effect never exceeds *|*0.04*|* PHQ-8 points anywhere on the grid, about one tenth of the food-insecurity gradient. The conclusion that the schooling channel is negligible therefore does not rest on any assumption about the strength of contemporaneous confounding. Early human capital thus reaches adult stature directly, while adult mental health is shaped chiefly by contemporaneous hardship.

**Table 9.** Mediation of the early-capital association through schooling (SEM paths).

|  | (1) |
| --- | --- |
| *Mediator equation: Grades completed (R7)* |  |
| Height-for-age z (R1) | 0.027<br>(0.028) |
| *Outcome equation: PHQ-8 adult (R7)* |  |
| Grades completed (R7) | −0.069***<br>(0.021) |
| Height-for-age z (R1) | 0.007<br>(0.044) |
| Observations | 5730 |

Structural equation model estimated by maximum likelihood; both equations include the predetermined covariates and country and cohort indicators. Observed-information-matrix standard errors in parentheses. \* $p < 0.10$ , \*\* $p < 0.05$ , \*\*\* $p < 0.01$ .

## Discussion

Three findings stand out. First, early human capital has a long reach into adult *physical* outcomes—one standard deviation of infant height-for-age translates into 1.6 cm of adult stature (the total association with the continuous score; 1.20 is the corresponding spline segment of Table 3), echoing the stature-status literature [7]—but not into adult mental health, suggesting that depressive and anxiety symptoms in early adulthood are shaped more by proximate stressors than by early nutritional endowments. This null is not an artefact of measurement error, since it survives the latent-factor correction in the spirit of Cunha et al. [5].

Second, household food insecurity during COVID-19 is robustly associated with worse adult mental health three to four years after the shock. The depression and anxiety results replicate each other, but they are not independent evidence: the two scores are strongly correlated in adulthood (*r* = 0.76), so the GAD-7 findings are best read as replication in a closely related instrument rather than as corroboration from a second source. The association works through the persistence of both distress and material hardship. This extends the contemporaneous evidence from the same cohorts [14, 15] and from other LMICs [16] to the medium run, and is consistent with cumulative-adversity models of mental health [21]: in every specification, the count of childhood shocks remains an independent correlate of adult symptoms.

Third, in exploratory analyses the gradient is patterned by context and life stage rather than by early endowments. Across countries, the gradient is steepest in India and flat in Ethiopia—an ordering that survives the wild cluster bootstrap and within-country standardisation, but whose substantive reading remains open: the same measured shock can carry very different mental-health consequences depending on the depth of the national crisis and coping options, yet differential item functioning across languages and floor effects would produce exactly the observed signature (lowest symptom level with steepest slope in India, highest food-insecurity prevalence with no gradient in Ethiopia), and item-level data to test measurement invariance are not available in the constructed files. Across cohorts, the gradient *appears larger* in the Older Cohort, which faced the pandemic at about age 26—in the thick of labour-market insertion and family formation—rather than at 19; the interaction is marginal, the cohort-specific intervals overlap, the contrast does not survive a Holm adjustment, and age at the shock cannot be separated from age at measurement. With those caveats, the life-stage pattern is consistent with developmental accounts of emerging adulthood [34] and with the view that the transition from adolescence to adulthood is a sensitive period for mental health in its own right [35]. Finally, and contrary to the dynamic-complementarity prediction [4], early human capital did not buffer the pandemic shock; the interval on the interaction rules out all but small buffering effects. Together these results shift the policy emphasis from early endowments as a source of resilience toward the protection of household food security when macro-shocks strike, with particular attention to young adults, who are typically outside the safety nets that reach schoolchildren.

Several limitations temper these conclusions. First and most fundamental, mental health was first measured in 2020, during the shock itself, so there is no pre-pandemic baseline: the medium-term gradient is a conditional association, and a stable individual propensity to report both hardship and distress can account for part of it—a benchmark calculation using the observed contemporaneous association and persistence implies roughly 0.24 of the 0.41 depression gradient, though because both inputs are attenuated by measurement error this is an illustrative benchmark rather than an upper bound, and the trait channel could be larger; the residual cannot be separated into causal effect and time-varying confounding with these data. Round 5 (2016) did not carry the PHQ-8 or GAD-7, and no pre-shock psychosocial measure is included in deposit SN 9543 (the round-specific Young Lives deposits, which do contain psychosocial scales, are outside this replication package), so no pre-shock proxy adjustment is possible here. Food insecurity is the only pandemic stressor retained in the constructed files. Round 7 was not fielded in Vietnam—by design rather than through attrition—and within the three remaining countries retention is non-random; selection on observables is addressed by inverse-probability weights built both from the Round-1 vector and, sequentially, from the Round-6 outcomes themselves, both leave the gradient essentially unchanged (Appendix Table A5), the Lee bounds for the binary exposure exclude zero, and the two-step bootstrap confirms the standard errors; selection on post-2020 unobservables orthogonal to the lagged outcome remains untestable. The value-added and interaction models are associational, and the mediation decomposition assumes sequential exogeneity, though the sensitivity analysis bounds the indirect effect at *|*0.04*|* points across error correlations spanning *±*0.9 (Appendix Table A6). The round-frequency persistence parameter is not identified: the Sargan test rejects every instrument configuration, which renders the GMM estimators inconsistent, so we draw no conclusion about round-to-round self-productivity in either direction (Appendix Table A4). The number of clusters (56–60) is adequate but moderate, and country-specific estimates rest on 16–20 sentinel sites each; wild-bootstrap inference is therefore reported for everything country-specific, and all heterogeneity is labelled exploratory. Finally, the Young Lives sentinel-site design is pro-poor and not nationally representative, so levels and gradients should not be read as national estimates. These caveats motivate, rather than undermine, the central message.

## Conclusion

Using a 22-year, multi-country cohort, we document two findings that hold to the strictest reading of the evidence and two that are exploratory. What holds: a persistent, precisely estimated conditional gradient links pandemic food insecurity to young-adult depression and anxiety three to four years on, travelling with the persistence of hardship and distress and robust to attrition corrections and bounds; and early human capital, despite its strong reach into adult stature, neither predicts adult mental health—a precise null—nor buffers the shock. What is exploratory: the gradient differs across countries and appears larger in the cohort hit as young adults. Safeguarding the mental health of young adults in LMICs will depend less on early endowments than on shielding households—and especially young adults in the transition to work and family life—from food insecurity when large shocks strike.

## Appendix

This appendix collects the specification tests and the robustness and heterogeneity analyses referenced in the text (Tables A1–A9).

**Table A1.** Specification and diagnostic tests.

| Test | Statistic | *p* |
| --- | --- | --- |
| *Baseline linear model for adult PHQ-8 (Table 4, column 1)* |  |  |
| Breusch–Pagan/Cook–Weisberg heteroskedasticity | $\chi^2(1) = 539.4$ | < 0.001 |
| Ramsey RESET (powers of fitted values) | $F(3, 5734) = 6.88$ | < 0.001 |
| Link test (squared linear prediction) | $t = 4.06$ | < 0.001 |
| Mean variance inflation factor | 1.91 (max 2.67) | – |
| *Interaction model (Table 6); joint tests use the cluster-robust VCE* |  |  |
| Breusch–Pagan/Cook–Weisberg heteroskedasticity | $\chi^2(1) = 888.0$ | < 0.001 |
| Ramsey RESET (powers of fitted values) | $F(3, 5112) = 18.53$ | < 0.001 |
| HAZ × food insecurity (resilience, P3) | $F(1, 55) = 1.19$ | 0.280 |
| Female × food insecurity | $F(1, 55) = 0.05$ | 0.828 |
| Country × food insecurity (joint) | $F(2, 55) = 4.85$ | 0.012 |
| Female × HAZ | $F(1, 55) = 0.23$ | 0.631 |
| All interactions (joint) | $F(5, 55) = 2.40$ | 0.049 |
| *Cohort interactions (Table A3, columns 1–2)* |  |  |
| Cohort × food insecurity, PHQ-8 | $F(1, 55) = 4.53$ | 0.038 |
| Cohort × food insecurity, GAD-7 | $F(1, 55) = 6.55$ | 0.013 |
| Cohort × HAZ, PHQ-8 | $F(1, 55) = 0.02$ | 0.887 |
| Cohort × HAZ, GAD-7 | $F(1, 55) = 1.30$ | 0.260 |
| *Formal-employment logit (Table 7, column 1)* |  |  |
| Hosmer–Lemeshow goodness of fit (10 groups) | $\chi^2(8) = 14.00$ | 0.082 |
| Area under the ROC curve | 0.729 | – |
| *Static and dynamic panel models (Table 8; baseline instruments)* |  |  |
| Hausman test, fixed versus random effects | $\chi^2(5) = 390.9$ | < 0.001 |
| Arellano–Bond AR(1) | $z = -16.12$ | < 0.001 |
| Arellano–Bond AR(2) | $z = 1.99$ | 0.047 |
| Sargan overidentification | $\chi^2(8) = 168.8$ | < 0.001 |
| *Overidentified measurement model (4 indicators, correlated Round-1 uniquenesses)* |  |  |
| Model versus saturated | $\chi^2(1) = 21.79$ | < 0.001 |
| RMSEA (90% CI) | 0.044 | – |
| Probability RMSEA $\leq 0.05$ | 0.704 | – |
| CFI / TLI | 0.999 / 0.994 | – |
| SRMR | 0.005 | – |

Linear-model diagnostics are computed on the homoskedastic version of each model, as the classical tests require; reported inference throughout the paper uses cluster-robust standard errors. Interaction and cohort tests are Wald tests on the cluster-robust variance matrix (55 residual degrees of freedom). Deep-lag AR and Sargan statistics for the dynamic panel are in Table A4. With $N = 10,831$ , the $\chi^2$ test of the measurement model is sensitive to trivial misfit; the approximation indices (RMSEA, CFI, TLI, SRMR) all indicate close fit.

**Table A2.** Country-specific estimates of the pandemic gradient (full models).

|  | PHQ-8 adult (R7) |  |  | GAD-7 adult (R7) |  |  |
| --- | --- | --- | --- | --- | --- | --- |
|  | (1)<br>Ethiopia | (2)<br>India | (3)<br>Peru | (4)<br>Ethiopia | (5)<br>India | (6)<br>Peru |
| Food insecurity, COVID (R6) | 0.002<br>(0.146) | 0.579***<br>(0.140) | 0.411**<br>(0.147) | -0.043<br>(0.167) | 0.597***<br>(0.110) | 0.418**<br>(0.177) |
| Height-for-age z (R1) | 0.079<br>(0.104) | -0.050<br>(0.055) | -0.204<br>(0.150) | 0.021<br>(0.096) | -0.039<br>(0.062) | -0.044<br>(0.121) |
| Early stunting (R1) | 0.175<br>(0.350) | -0.123<br>(0.167) | -0.375<br>(0.398) | -0.036<br>(0.343) | -0.083<br>(0.179) | -0.107<br>(0.317) |
| Housing quality (R1) | 0.561<br>(0.511) | -0.503*<br>(0.277) | -0.335<br>(0.404) | 0.433<br>(0.737) | -0.265<br>(0.300) | 0.104<br>(0.527) |
| Services access (R1) | -0.111<br>(0.431) | -0.180<br>(0.494) | 0.464<br>(0.501) | -0.059<br>(0.358) | -0.477<br>(0.412) | 0.270<br>(0.529) |
| Consumer durables (R1) | 0.295<br>(0.708) | -0.515<br>(0.351) | 1.482**<br>(0.700) | -0.879<br>(0.962) | -0.450*<br>(0.231) | 1.603**<br>(0.703) |
| Mother's education (grades) | -0.002<br>(0.016) | -0.019<br>(0.021) | 0.012<br>(0.029) | -0.012<br>(0.017) | -0.027<br>(0.020) | -0.008<br>(0.031) |
| Father's education (grades) | 0.002<br>(0.014) | 0.017<br>(0.012) | 0.022<br>(0.035) | 0.010<br>(0.013) | 0.007<br>(0.010) | -0.024<br>(0.037) |
| Female | 0.450<br>(0.267) | 0.336*<br>(0.175) | 1.491***<br>(0.179) | 0.492***<br>(0.146) | 0.493**<br>(0.215) | 1.942***<br>(0.207) |
| Urban | 0.734*<br>(0.411) | 0.086<br>(0.304) | 0.365<br>(0.238) | 1.121***<br>(0.340) | 0.500*<br>(0.282) | 0.646***<br>(0.197) |
| Cumulative household shocks (0-5) | 0.393**<br>(0.135) | 0.172**<br>(0.076) | 0.212***<br>(0.066) | 0.423***<br>(0.077) | 0.149**<br>(0.066) | 0.299***<br>(0.067) |
| Younger cohort | -0.313<br>(0.277) | -0.111<br>(0.175) | 0.834***<br>(0.234) | -0.353<br>(0.326) | -0.311*<br>(0.154) | 0.144<br>(0.267) |
| Constant | -0.433<br>(0.835) | 0.125<br>(0.459) | -0.632<br>(0.515) | -0.359<br>(0.365) | 0.433<br>(0.395) | 0.358<br>(0.454) |
| Observations | 1113 | 2512 | 1510 | 1115 | 2497 | 1520 |
| Adjusted R-squared | 0.031 | 0.041 | 0.055 | 0.043 | 0.045 | 0.069 |

Each column estimates Eq (2) on one country's subsample (country fixed effects therefore drop out). Cluster-robust standard errors in parentheses (16 sentinel-site clusters in Ethiopia, 20 in India, 20 in Peru); given the small cluster counts, the food-insecurity coefficients are re-evaluated with the wild cluster bootstrap- $t$ (Webb weights, 999 replications, null imposed): $p_{\text{wild}} = 0.98, 0.001, 0.013$ for PHQ-8 and $0.80, < 0.001, 0.033$ for GAD-7 in Ethiopia, India, and Peru respectively. With outcome and exposure standardised within country, the gradients are $0.000, 0.120, 0.061$ (PHQ-8) and $-0.009, 0.123, 0.058$ (GAD-7) standard deviations. These contrasts are exploratory. \* $p < 0.10$ , \*\* $p < 0.05$ , \*\*\* $p < 0.01$ .

**Table A3.** Cohort heterogeneity of the pandemic gradient: interaction and subsample models.

|  | Interaction models |  | Cohort subsamples |  |  |  |
| --- | --- | --- | --- | --- | --- | --- |
|  | (1)<br>PHQ-8 | (2)<br>GAD-7 | (3)<br>PHQ-8, 22 | (4)<br>PHQ-8, 29 | (5)<br>GAD-7, 22 | (6)<br>GAD-7, 29 |
| Food insecurity, COVID (R6) | 0.746***<br>(0.202) | 0.794***<br>(0.200) | 0.327***<br>(0.101) | 0.574**<br>(0.217) | 0.290***<br>(0.094) | 0.717***<br>(0.210) |
| Height-for-age z (R1) | −0.050<br>(0.095) | 0.056<br>(0.080) | −0.050<br>(0.057) | 0.006<br>(0.122) | −0.029<br>(0.051) | 0.058<br>(0.128) |
| Younger cohort | 0.958***<br>(0.345) | 0.632**<br>(0.301) |  |  |  |  |
| Younger × Food insecurity (R6) | −0.471**<br>(0.221) | −0.526**<br>(0.206) |  |  |  |  |
| Younger × HAZ (R1) | 0.012<br>(0.081) | −0.085<br>(0.075) |  |  |  |  |
| Female | 0.716***<br>(0.134) | 0.944***<br>(0.152) | 0.655***<br>(0.143) | 0.855***<br>(0.194) | 0.868***<br>(0.164) | 1.128***<br>(0.225) |
| Cumulative shocks | 0.257***<br>(0.046) | 0.287***<br>(0.045) | 0.257***<br>(0.045) | 0.266***<br>(0.082) | 0.271***<br>(0.049) | 0.329***<br>(0.090) |
| Observations | 5135 | 5132 | 3707 | 1428 | 3704 | 1428 |
| Adjusted R-squared | 0.096 | 0.133 | 0.108 | 0.066 | 0.138 | 0.123 |

All models include the full predetermined covariate vector and country fixed effects; columns 1–2 additionally interact cohort with the shock and with early height-for-age. “22” and “29” denote the Younger and Older Cohort subsamples (ages at Round 7). Cluster-robust standard errors in parentheses (56 sentinel-site clusters in columns 1–2; 54–56 in columns 3–6). The cohort contrasts are exploratory: the interaction $p$ values (0.038 and 0.013) do not survive a Holm adjustment within the family of heterogeneity tests, the subsample intervals overlap, and age at the shock, age at measurement, and birth cohort are perfectly collinear in this design. \* $p < 0.10$ , \*\* $p < 0.05$ , \*\*\* $p < 0.01$ .

**Table A4.** Dynamic panel robustness: restricting the GMM instrument set to deep lags.

|  | (1)<br>Baseline (lags 2+) | (2)<br>Deep lags (3+) | (3)<br>Single lag 3 |
| --- | --- | --- | --- |
| Lagged height-for-age z | 0.191***<br>(0.013) | −0.015<br>(0.052) | −0.010<br>(0.053) |
| Wealth index | 0.398***<br>(0.051) | 0.455***<br>(0.046) | 0.455***<br>(0.046) |
| Number of instruments | 8 | 4 | 3 |
| Arellano–Bond AR(1), $p$ | < 0.001 | < 0.001 | < 0.001 |
| Arellano–Bond AR(2), $p$ | 0.047 | 0.172 | 0.223 |
| Sargan test, $p$ | < 0.001 | < 0.001 | < 0.001 |
| Observations | 16527 | 16527 | 16527 |

**Table A5.** Attrition: retention probits and inverse-probability-weighted estimates, including sequential weights.

|  | (1)<br>Retention | (2)<br>Retention, seq. | (3)<br>PHQ-8 | (4)<br>PHQ-8, IPW | (5)<br>PHQ-8, seq. | (6)<br>GAD-7, IPW | (7)<br>GAD-7, seq. |
| --- | --- | --- | --- | --- | --- | --- | --- |
| Food insecurity, COVID (R6) |  | −0.267***<br>(0.086) | 0.407***<br>(0.093) | 0.381***<br>(0.091) | 0.402***<br>(0.093) | 0.366***<br>(0.100) | 0.404***<br>(0.099) |
| PHQ-8 during COVID (R6) |  | −0.018<br>(0.028) |  |  |  |  |  |
| GAD-7 during COVID (R6) |  | −0.005<br>(0.028) |  |  |  |  |  |
| Height-for-age z (R1) | 0.007<br>(0.020) | 0.046<br>(0.037) | −0.040<br>(0.051) | −0.037<br>(0.052) | −0.038<br>(0.051) | −0.024<br>(0.048) | −0.020<br>(0.047) |
| Early stunting (R1) | −0.032<br>(0.055) | 0.082<br>(0.119) | −0.147<br>(0.155) | −0.140<br>(0.159) | −0.143<br>(0.155) | −0.120<br>(0.151) | −0.104<br>(0.145) |
| Female | 0.082**<br>(0.040) | 0.039<br>(0.078) | 0.711***<br>(0.135) | 0.708***<br>(0.134) | 0.707***<br>(0.134) | 0.924***<br>(0.146) | 0.928***<br>(0.149) |
| Cumulative shocks | 0.270***<br>(0.030) | −0.006<br>(0.042) | 0.260***<br>(0.047) | 0.266***<br>(0.046) | 0.264***<br>(0.047) | 0.292***<br>(0.044) | 0.283***<br>(0.045) |
| Observations | 7228 | 5252 | 5135 | 5135 | 5134 | 5074 | 5073 |
| Adjusted R-squared |  |  | 0.095 | 0.094 | 0.095 | 0.132 | 0.132 |
| Pseudo R-squared | 0.101 | 0.061 |  |  |  |  |  |

Column 1: probit of being observed for adult PHQ-8 at Round 7 on the predetermined Round-1 vector (Ethiopia, India, and Peru; Vietnam is excluded because Round 7 was not fielded there by design). Column 2: sequential retention probit on the Round-6 sample, augmenting the Round-1 vector with the Round-6 outcomes themselves; pandemic-period symptoms do not predict drop-out. Columns 3–7 re-estimate the COVID specification of Eq (2) unweighted, with Round-1 inverse-probability weights, and with sequential weights (both sets trimmed at the 99th percentile). All models show selected coefficients; the full predetermined vector, cohort, and country fixed effects are included throughout. Cluster-robust standard errors in parentheses (56–60 sentinel-site clusters). \* $p < 0.10$ , \*\* $p < 0.05$ , \*\*\* $p < 0.01$ .

**Table A6.** Sensitivity of the mediation decomposition to correlated errors over the near-full feasible range.

| | Assumed error correlation $\rho$ between Eqs (9) and (10) | | | | | | |
| --- | --- | --- | --- | --- | --- | --- | --- |
|  | −0.9 | −0.6 | −0.3 | 0 | 0.3 | 0.6 | 0.9 |
| $a$ (HAZ → schooling) | 0.027 | 0.027 | 0.027 | 0.027 | 0.027 | 0.027 | 0.027 |
| $b$ (schooling → PHQ-8) | 1.330 | 0.864 | 0.397 | −0.069 | −0.536 | −1.002 | −1.469 |
| Indirect effect $ab$ | 0.036 | 0.023 | 0.011 | −0.002 | −0.014 | −0.027 | −0.039 |
| Direct effect $c'$ | −0.030 | −0.018 | −0.005 | 0.007 | 0.020 | 0.032 | 0.045 |

Each column re-estimates the system of Eqs (9)–(10) by maximum likelihood with the error covariance fixed at $\rho\sigma_\xi\sigma_\zeta$ (baseline error variances 4.27 and 10.32; $N = 5,730$ ; all constrained models converge). The grid spans essentially the entire feasible range of contemporaneous confounding, so the exercise is not conditioned on any assumption about its strength. The coefficient $a$ is invariant to $\rho$ by construction, and it is this invariant, near-zero first stage—not the grid—that bounds the indirect effect: although $b$ is unidentified under simultaneity (it swings from +1.33 to −1.47), $ab$ never exceeds $|0.04|$ PHQ-8 points, about one tenth of the food-insecurity gradient.

**Table A7.** Ordinal coding of the pandemic food-insecurity exposure.

|  | (1)<br>PHQ-8 | (2)<br>GAD-7 |
| --- | --- | --- |
| Food insecurity = 2 (vs 1) | 0.303**<br>(0.118) | 0.273**<br>(0.112) |
| Food insecurity = 3 (vs 1) | 1.082***<br>(0.260) | 1.129***<br>(0.274) |
| Food insecurity = 4 (vs 1) | 0.292<br>(0.809) | 0.550<br>(0.721) |
| Height-for-age z (R1) | −0.040<br>(0.051) | −0.019<br>(0.047) |
| Observations | 5135 | 5132 |
| Adjusted R-squared | 0.095 | 0.133 |

Eq (2) with the linear food-insecurity term replaced by category indicators (base: 1 = always eats enough; 2 = sometimes short; 3 = often short; 4 = frequently does not eat enough). The response is monotone over the well-populated range; category 4 is rare and imprecisely estimated. Wald tests of the equidistance restriction ( $b_3 = 2b_2$ and $b_4 = 3b_2$ ) do not reject ( $p = 0.19$ for PHQ-8 and $p = 0.12$ for GAD-7), but because the top category contributes almost no information, this non-rejection reflects limited power against alternatives involving that category as much as support for linearity; the linear term is best read as an average gradient over the well-populated range. Full predetermined vector, cohort, and country fixed effects included; cluster-robust standard errors in parentheses (56 sentinel-site clusters). \* $p < 0.10$ , \*\* $p < 0.05$ , \*\*\* $p < 0.01$ .

**Table A8.** Persistence chain: each link conditions only on predetermined covariates and its own lag.

|  | (1)<br>Food insecurity (R7) | (2)<br>PHQ-8 (R7) | (3)<br>GAD-7 (R7) |
| --- | --- | --- | --- |
| Food insecurity, COVID (R6) | 0.191***<br>(0.018) |  |  |
| PHQ-8 during COVID (R6) |  | 0.412***<br>(0.041) |  |
| GAD-7 during COVID (R6) |  |  | 0.467***<br>(0.035) |
| Height-for-age z (R1) | −0.018***<br>(0.007) | 0.011<br>(0.050) | 0.042<br>(0.052) |
| Observations | 5258 | 5572 | 5564 |
| Adjusted R-squared | 0.170 | 0.145 | 0.193 |

Collider-free characterisation of the channels behind the value-added decomposition: each regression conditions only on the predetermined vector (with cohort and country fixed effects) and a single lagged variable, so no post-treatment variables are held fixed simultaneously. Pandemic food insecurity strongly predicts adult food insecurity, and each pandemic-period symptom score strongly predicts its adult counterpart. Cluster-robust standard errors in parentheses (56 sentinel-site clusters). \* $p < 0.10$ , \*\* $p < 0.05$ , \*\*\* $p < 0.01$ .

**Table A9.** Bounded-count and clinical-threshold models of the pandemic gradient.

| | (1)<br>PHQ-8 (Poisson) | (2)<br>GAD-7 (Poisson) | (3)<br>PHQ-8 $\geq 10$ (logit) | (4)<br>GAD-7 $\geq 10$ (logit) |
| --- | --- | --- | --- | --- |
| Food insecurity, COVID (R6) | 1.203***<br>(0.051) | 1.178***<br>(0.044) | 1.290*<br>(0.172) | 1.526***<br>(0.181) |
| Height-for-age z (R1) | 0.982<br>(0.027) | 0.994<br>(0.023) | 0.957<br>(0.100) | 0.965<br>(0.085) |
| Female | 1.418***<br>(0.082) | 1.501***<br>(0.083) | 2.478***<br>(0.373) | 2.888***<br>(0.393) |
| Cumulative shocks | 1.124***<br>(0.026) | 1.117***<br>(0.021) | 1.178**<br>(0.077) | 1.241***<br>(0.071) |
| Observations | 5135 | 5132 | 5135 | 5132 |

The symptom scores are bounded counts with a large mass at zero (base rates of the clinical thresholds: 5.6% for PHQ-8 $\geq 10$ and 7.6% for GAD-7 $\geq 10$ ). Columns 1–2: Poisson quasi-maximum-likelihood incidence-rate ratios; the average marginal effect of one step of pandemic food insecurity is +0.37 PHQ-8 points and +0.40 GAD-7 points. Columns 3–4: odds ratios for exceeding the clinical threshold; the average marginal effects are +1.0 percentage point for PHQ-8 $\geq 10$ ( $p = 0.07$ ) and +1.9 percentage points for GAD-7 $\geq 10$ ( $p = 0.001$ ) per step. Full predetermined vector, cohort, and country fixed effects included; exponentiated coefficients with cluster-robust standard errors in parentheses (56 sentinel-site clusters). \* $p < 0.10$ , \*\* $p < 0.05$ , \*\*\* $p < 0.01$ .

## Author contributions

Conceptualization: all authors. Methodology: MADF, WMJR. Formal analysis: MADF, WMJR. Data curation: WMJR, GMPH. Validation: GMPH, URM. Visualization: MADF, URM. Writing—original draft: MADF, GMPH. Writing—review and editing: all authors. (CRediT taxonomy; contributions are also entered per author in the submission system.)

## Competing interests

The authors have declared that no competing interests exist.

## Data availability

The data underlying this study are third-party data: the Young Lives Rounds 1–7 constructed files, available from the UK Data Service (study number SN 9543). Access requires free registration with the UK Data Service and acceptance of its End User Licence; the authors accessed the data under those standard conditions and had no special access privileges that other researchers would not have. The single Stata do-file that reproduces every number, table, and figure in this manuscript from those files is provided as S1 File.

## Acknowledgments

The data used in this publication come from Young Lives, a 20-year study of childhood poverty and transitions to adulthood in Ethiopia, India, Peru, and Vietnam (www.younglives.org.uk), accessed through the UK Data Service (study number 9543). Young Lives is funded by UK aid from the Foreign, Commonwealth & Development Office and a number of further funders. The views expressed here are those of the authors; they are not necessarily those of Young Lives, the University of Oxford, or the funders.

## Supporting information

**S1 File. Stata replication code.** A single, self-contained Stata do-file (young lives plos master.do) that constructs the analysis files from the UK Data Service constructed datasets (study 9543) and reproduces every number, table, and figure reported in this manuscript in one run.

**S2 Checklist. STROBE checklist.** Completed STROBE checklist for observational (cohort) studies, mapping each item to the corresponding manuscript section.

## Notes

### Competing Interest Statement

The authors have declared no competing interest.

### Author Declarations

This study is a secondary analysis of fully anonymised, publicly archived data collected by the Young Lives study, which obtained ethical approval from the University of Oxford Central University Research Ethics Committee and from country-level committees. Informed consent—written or, where literacy was limited, documented oral consent—was obtained from caregivers and participants at every round. The anonymised data were accessed through the UK Data Service (study number 9543) no additional ethical approval was required for this analysis.

